# Hospital Waste Management Readiness in Urban Bangladesh: A Knowledge, Attitudes, and Practices Assessment

**DOI:** 10.64898/2026.05.25.26354076

**Authors:** Nur Naher Bhuiyan, Kamrun Naher Bhuiyan, Sarmin Aktar, Rajat Sanker Roy Biswas, Tofazzal Md Rakib, Mohammad Alamgir Hossain

## Abstract

Healthcare waste (HCW) management is a critical determinant of occupational safety, infection control, and environmental protection, particularly in low- and middle-income settings. Using the knowledge–attitude–practice (KAP) framework, this study assessed cognitive, behavioral, and institutional dimensions of HCW management among healthcare workers in urban Bangladesh. A cross-sectional survey was conducted among 342 cleaners and nurses in hospitals in the Chattogram Metropolitan Area (CMA) and Cumilla City Corporation (CuCC). Marked disparities were observed across professional groups. Training coverage was significantly lower among nurses than cleaners in CMA (22.5% vs. 48.7%; p = 0.002), whereas in CuCC nurses showed higher coverage (69.0% vs. 52.3%; p < 0.01). Knowledge of color-coded waste segregation was generally inadequate, with only 39.3% of CMA cleaners correctly identifying pharmaceutical waste bins compared with 60.0% of nurses (p < 0.01); CuCC nurses demonstrated substantially higher awareness (82.8%). Attitudinal indicators favored nurses, with strong hygiene and environmental risk awareness (95–100%) compared with cleaners (66–87.3%; p < 0.001). Despite this, compliance with segregation practices remained low across both sites (<30%). Several institutional support indicators were more favorable among nurses, particularly in CuCC. These findings indicate a significant knowledge–practice gap, emphasizing that effective HCW management requires not only training but also strengthened institutional structures and enforcement mechanisms to reduce public health and environmental risks.

## 1. Introduction

Healthcare waste management (HCWM) remains a persistent global challenge and a critical component of healthcare system governance. Healthcare facilities worldwide depend on structured waste management systems to comply with regulatory standards and to ensure the safe handling of waste generated throughout clinical activities, from segregation and temporary storage to treatment and final disposal (Mathur et al., 2011; Chartier, 2014; Dang et al., 2021). Effective HCWM is essential for minimizing occupational hazards, preventing disease transmission, and protecting environmental health.

Healthcare waste (HCW) encompasses all waste produced during the diagnosis, treatment, or immunization of humans or animals, as well as related research and laboratory activities (Khan et al., 2019). This includes sharps, blood and body fluids, pharmaceuticals, chemicals, radioactive materials, and contaminated medical devices (Kagoma et al., 2012). HCW is commonly categorized into general (non-hazardous), infectious, hazardous, and radioactive waste (Kagoma et al., 2012; Malsparo, 2015). Although 75–85% of HCW is non-hazardous, the remaining 15–25% poses significant health and environmental risks if improperly managed (Janik-Karpinska et al., 2023).

Globally, the scale of HCW generation continues to increase with expanding healthcare services. The World Health Organization estimates that approximately 16 billion injections are administered annually, generating substantial volumes of waste requiring safe disposal (WHO, 2024). During the COVID-19 pandemic, medical waste surged dramatically in several Asian cities, highlighting the vulnerability of existing waste management systems (Jafarzadeh & Maleki, 2023). In Bangladesh, the Chattogram Metropolitan Area alone generates an estimated 15–18 tons of HCW per day, accounting for approximately 1–1.2% of municipal solid waste (Alam et al., 2013; Alam et al., 2015).

While high-income countries have established enforceable HCWM policies supported by adequate infrastructure and resources, many low- and middle-income countries struggle with implementation due to financial, technical, and institutional constraints (Mohee, 2005; Tudor et al., 2005). As a result, unsafe practices such as poor segregation, open dumping, open-air burning, and mixing HCW with municipal waste remain widespread (Mohankumar & Kottaiveeran, 2011; Kerdsuwan & Laohalidanond, 2015). These practices contribute to environmental contamination and increase the risk of occupational exposure, needle-stick injuries, and transmission of blood-borne pathogens such as hepatitis B, hepatitis C, and HIV (Vandana et al., 2020).

Effective HCWM relies heavily on proper waste segregation at the point of generation, typically through color-coded systems (Adogu et al., 2014). However, evidence from multiple settings indicates persistent failures in segregation, leading to increased treatment costs and environmental risks (Kumar et al., 2018). Studies further suggest that gaps in healthcare workers’ knowledge, attitudes, and practices (KAP), compounded by limited institutional support and weak monitoring, undermine compliance even where awareness exists (Devi et al., 2019; Rajpal et al., 2024). These challenges underscore the need for integrated HCWM approaches that align individual competencies with organizational capacity and regulatory enforcement.

This study assessed the knowledge, attitudes, and practices (KAP) related to healthcare waste management among nurses and cleaners in hospitals in the Chattogram Metropolitan Area and Cumilla City Corporation, Bangladesh, and examined professional and institutional factors influencing compliance with standard waste segregation and handling procedures. This study aimed to investigate whether disparities exist in training and knowledge between professional groups and reveal a persistent gap between positive attitudes and actual waste management practices, largely attributable to inadequate institutional support and monitoring.

## 2. Materials and Methods

### 2.1. Study design and setting

An observational cross-sectional study was conducted to assess the current status and patterns of healthcare waste (HCW) management practices in selected hospitals, clinics, and diagnostic laboratories located in the Chattogram Metropolitan Area (CMA) and Cumilla City Corporation (CuCC), Bangladesh. The study was carried out over an 18-month period from 1 January 2024 to 30 April 2025. Both government and private healthcare facilities were included to ensure representation of different healthcare service settings. Eligible participants included nurses and cleaning staff who had at least six months of work experience in their respective facilities. Healthcare workers who declined to provide informed consent were excluded from the study.

### 2.2. Sampling technique

A non-probability convenience and purposive sampling approach was employed. Selected healthcare facilities in CMA and CuCC were approached after obtaining institutional permission from the relevant authorities. Healthcare workers were recruited directly from their respective workplaces during routine duty hours. Nurses and cleaners who met the eligibility criteria were informed about the objectives and procedures of the study and were invited to participate voluntarily. Sampling was conducted proportionately across the selected facilities based on the availability of eligible staff during the study period. Participation was entirely voluntary, and eligible healthcare workers were given the option to accept or decline participation without any consequences. A total of 342 participants consented and were enrolled in the study, including 190 respondents from CMA and 152 from CuCC.

### 2.3. Data collection instrument and procedure

Data was collected using a structured, pre-tested questionnaire designed to assess knowledge, attitudes, and practices (KAP) related to HCW management. The questionnaire comprised sections on socio-demographic characteristics, occupational background, health status, work schedules, and perceived challenges related to waste management. The knowledge section evaluated participants’ understanding of HCW categories, color-coded segregation systems, and appropriate disposal methods for infectious, sharp, pharmaceutical, and cytotoxic waste. The attitude section assessed perceptions regarding the importance of HCW management, use of personal protective equipment (PPE), and adequacy of institutional support, including hygiene facilities and health monitoring. The practice section examined routine waste-handling behaviors, compliance with segregation protocols, PPE usage, and adherence to safety measures.

Data collection was conducted by trained research personnel through face-to-face interviews at the workplaces of respondents. Before questionnaire administration, participants received a brief explanation regarding the purpose of the study, confidentiality of information, and their right to withdraw at any stage or skip any question without penalty. Each interview required approximately 10–12 minutes to complete. Confidentiality and anonymity were strictly maintained throughout the study, and no invasive procedures or interventions were involved.

### 2.4. Data analysis

Data were initially entered, cleaned, and validated using Microsoft Excel 2016 (Microsoft Corp., Redmond, WA, USA) and subsequently exported to STATA SE version 18 (StataCorp, College Station, TX, USA) for statistical analysis. Descriptive statistics, including frequencies, percentages, means, and standard deviations, were used to summarize socio-demographic characteristics and knowledge, attitude, and practice (KAP) responses related to healthcare waste management. Associations between categorical variables, including professional category (cleaner vs. nurse) and study area (CMA vs. CuCC), were assessed using chi-square tests or Fisher’s exact tests where appropriate. A p-value of ≤ 0.05 was considered statistically significant.

## 3. Results

### 3.1. Participant characteristics

A total of 342 healthcare workers participated in the study, comprising 283 cleaners (80.4%) and 69 nurses (19.6%). Of the cleaners, 150 (53.0%) were recruited from the Chattogram Metropolitan Area (CMA) and 133 (47.0%) from Cumilla City Corporation (CuCC). Among nurses, 40 (57.9%) were from CMA and 29 (42.1%) from CuCC (Table 1). Female workers predominated overall, accounting for 57.2% of cleaners and 91.3% of nurses. All male nurses (n = 6) were recruited from CMA, whereas male cleaners were more frequently observed in CMA than CuCC (64.5% vs. 35.5%). Cleaners were generally older and had longer work experience than nurses across both locations. The mean age of cleaners was 35.3 ± 8.5 years in CMA and 40.2 ± 3.1 years in CuCC, compared with 30.1 ± 3.0 years and 33.2 ± 3.5 years among nurses, respectively. Mean duration of employment among cleaners exceeded five years in both CMA (64.1 ± 52.3 months) and CuCC (71.2 ± 16.8 months), whereas nurses reported shorter service duration (45.9 ± 19.5 months in CMA; 41.3 ± 17.3 months in CuCC) (Table 1).

**Table 1.** Demographic overview of the participants.

| Category | Sub-category | Healthcare worker type | CMA, n (%) | CuCC, n (%) | Total |
| --- | --- | --- | --- | --- | --- |
| Number of participants |  | Cleaner | 150 (53.0%) | 133 (47.0%) | 283 |
|  |  | Nurse | 40 (57.9%) | 29 (43.1%) | 69 |
| **Gender** | Female | Cleaner | 72 (44.4%) | 90 (56.6%) | 162 |
|  |  | Nurse | 34 (53.9%) | 29 (46.1%) | 63 |
|  | Male | Cleaner | 78 (64.5%) | 43 (35.5%) | 121 |
|  |  | Nurse | 6 (100%) | 0 | 6 |
| **Age (in years)**<br>**mean±SD)** |  | Cleaner | 35.3±8.5 | 40.2±3.1 |  |
|  |  | Nurse | 30.1±3.0 | 33.2±3.5 |  |
| **Experience (in**<br>**months; mean±SD)** |  | Cleaner | 64.1±52.3 | 71.2±16.8 |  |
|  |  | Nurse | 45.9±19.5 | 41.3±17.3 |  |

### 3.2. Knowledge of healthcare waste management

Substantial variation in HCW-related knowledge was observed across professional groups and locations. In CMA, a significantly lower proportion of nurses reported having received formal training on HCW management compared with cleaners (22.5% vs. 48.7%; p < 0.01). In CuCC, training coverage was higher among nurses than cleaners (69.0% vs. 52.3%), although the difference did not reach statistical significance. Training coverage differed between locations within worker categories, particularly among nurses (Table 2).

**Table 2.** Training status regarding HCW management in CMA and CuCC.

| Location | Worker type | Yes, n (%) | *p*-value†<br>(Cleaner vs<br>Nurse) | *p*-value‡<br>(Overall) |
| --- | --- | --- | --- | --- |
| **CMA** | Cleaner | 73 (48.7%) | **0.002** | **0.04** |
|  | Nurse | 9 (22.5%) |  |  |
| **CuCC** | Cleaner | 70 (52.3%) | 0.1 |  |
|  | Nurse | 20 (69%) |  |  |

† Chi-square test comparing Cleaner vs Nurse within each location.
‡ Chi-square test comparing CMA vs CuCC within each worker type.

Knowledge of color-coded waste segregation was inconsistent. In CMA, nurses demonstrated significantly better knowledge than cleaners regarding pharmaceutical waste segregation (60.0% vs. 39.3%; p < 0.01) and the importance of separate packaging (p < 0.05), while knowledge of infectious, chemical, and biological waste did not differ significantly (Table 3). In CuCC, nurses consistently outperformed cleaners across all waste categories, including infectious, chemical, pharmaceutical, and biological waste, as well as awareness of separate packaging (all p < 0.01). When pooled, overall knowledge did not differ significantly by location, indicating that professional role rather than geography accounted for most observed differences.

**Table 3.**
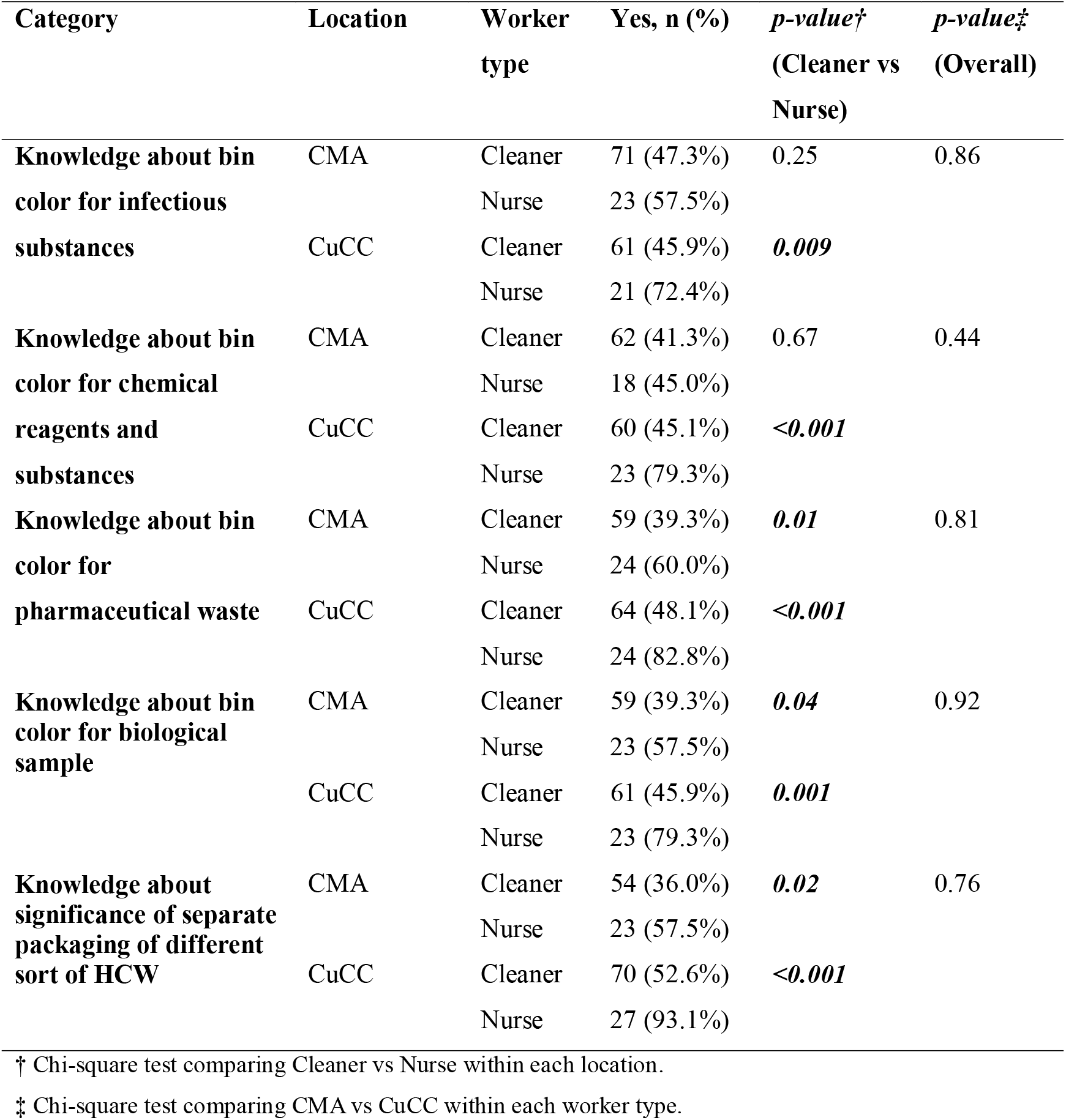
Knowledge regarding the use of color-coded bins for discarding HCW.

### 3.3. Attitudes toward HCW management

Attitudinal indicators were generally positive, particularly among nurses (Figure 1). Awareness of personal hygiene was universal among nurses in both locations and significantly higher than among cleaners in CMA (87.3%) and CuCC (86.5%) (p < 0.05). Awareness of environmental risks associated with HCW was also higher among nurses than cleaners in both CMA (95.0% vs. 66.0%; p < 0.001) and CuCC (100% vs. 83.5%; p < 0.01). Willingness to attend HCW training was high overall, with nurses in CMA showing significantly greater interest than cleaners (90.0% vs. 70.6%; p < 0.01). A significant location-wise difference in training willingness was also observed (p < 0.001).

**Figure 1.**
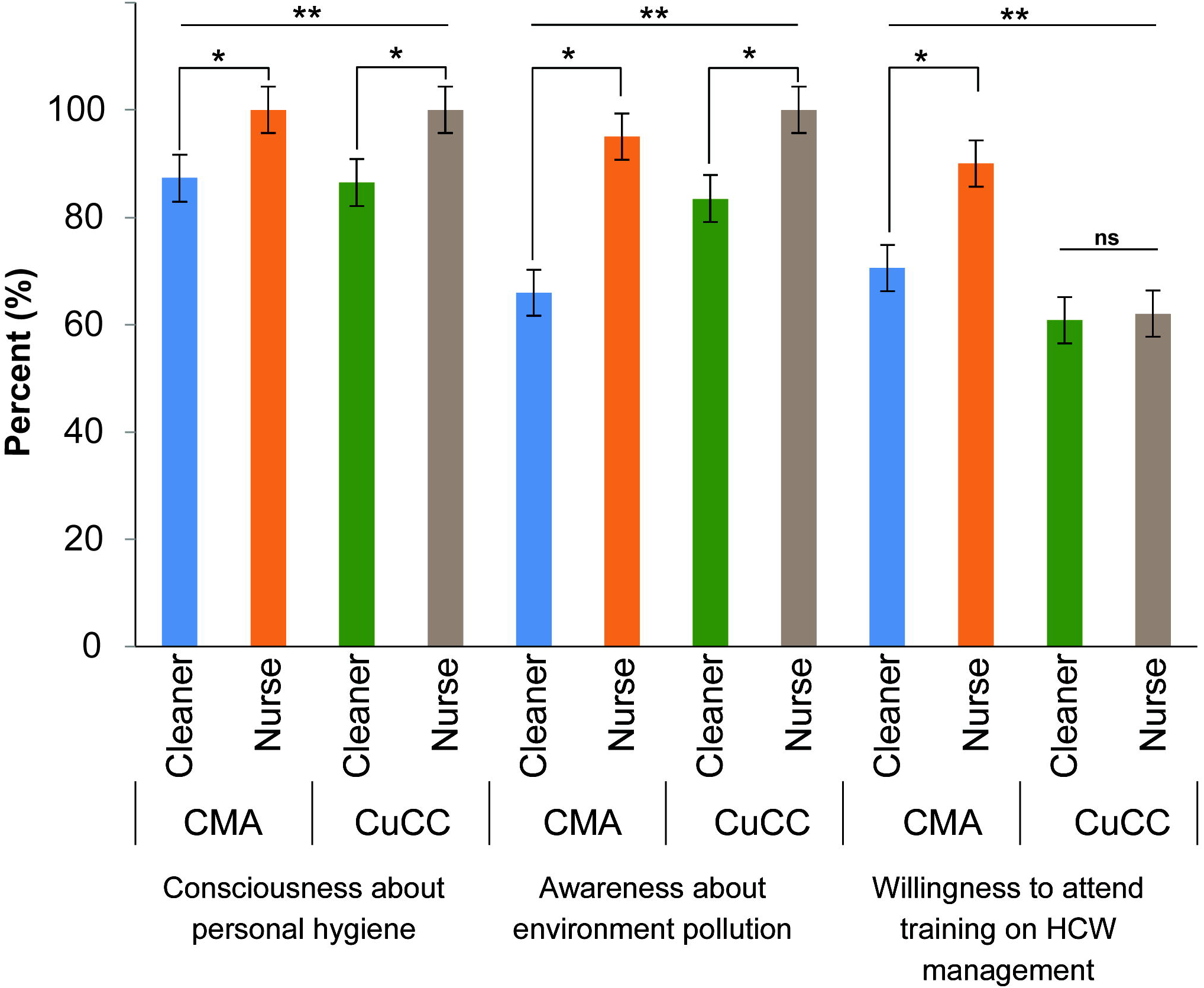
Attitudinal indicators related to healthcare waste (HCW) management among cleaners and nurses at CMA and CuCC. Percentages represent respondents’ awareness and attitudes regarding personal hygiene, environmental pollution, and willingness to participate in HCW management training. Asterisks indicate statistically significant differences based on chi-square tests: *P* < 0.05 for comparisons between cleaners and nurses within the same location (*), and between CMA and CuCC within the same worker category (**). “ns” indicates no statistically significant difference.

### 3.4. HCW management practices

Despite relatively favorable attitudes, adherence to recommended practices remained limited (Figure 2 and Table 4). Use of color-coded bins was particularly low in CMA, where only 19.3% of cleaners reported compliance and no nurses reported using the system (p < 0.001). In CuCC, compliance was modestly higher among both cleaners (28.6%) and nurses (24.1%), with a significant overall difference by location (p < 0.05) (Figure 2).

**Table 4.**
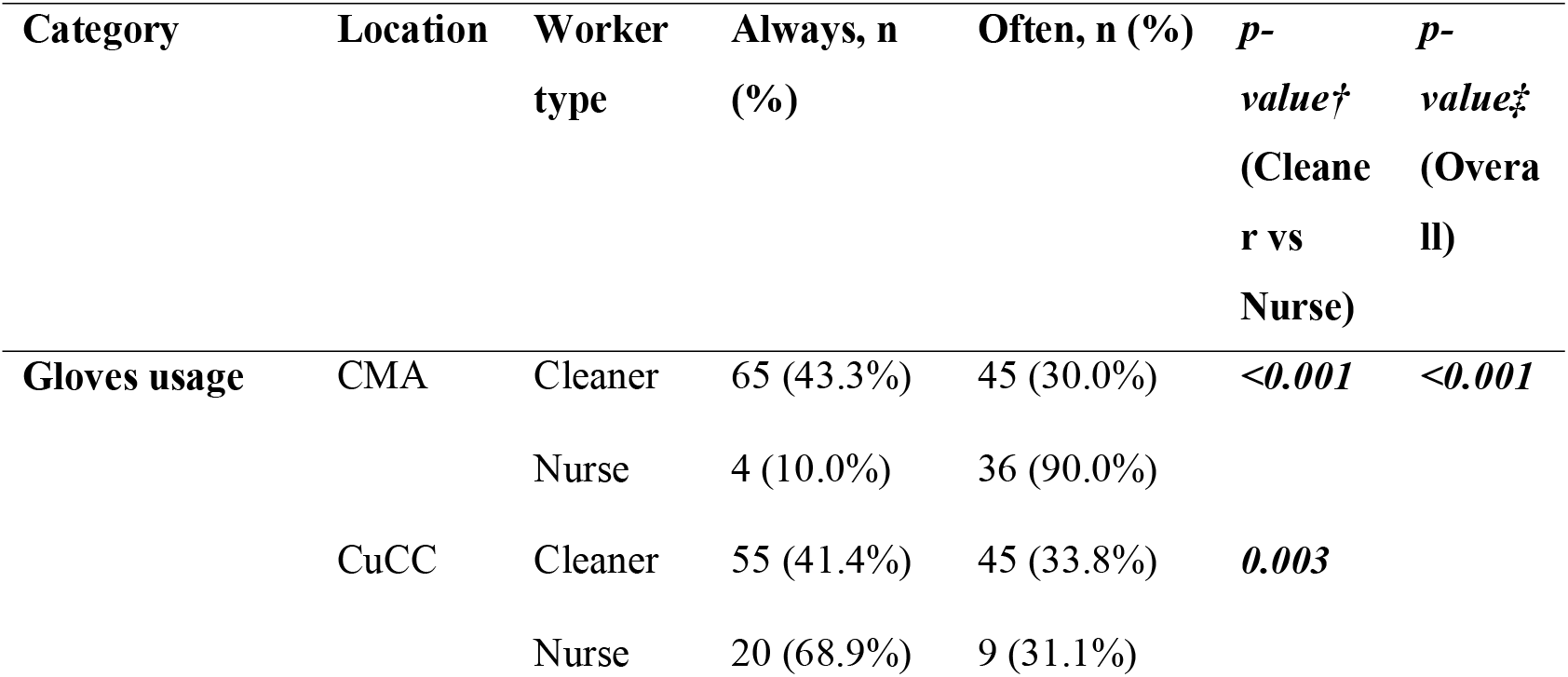
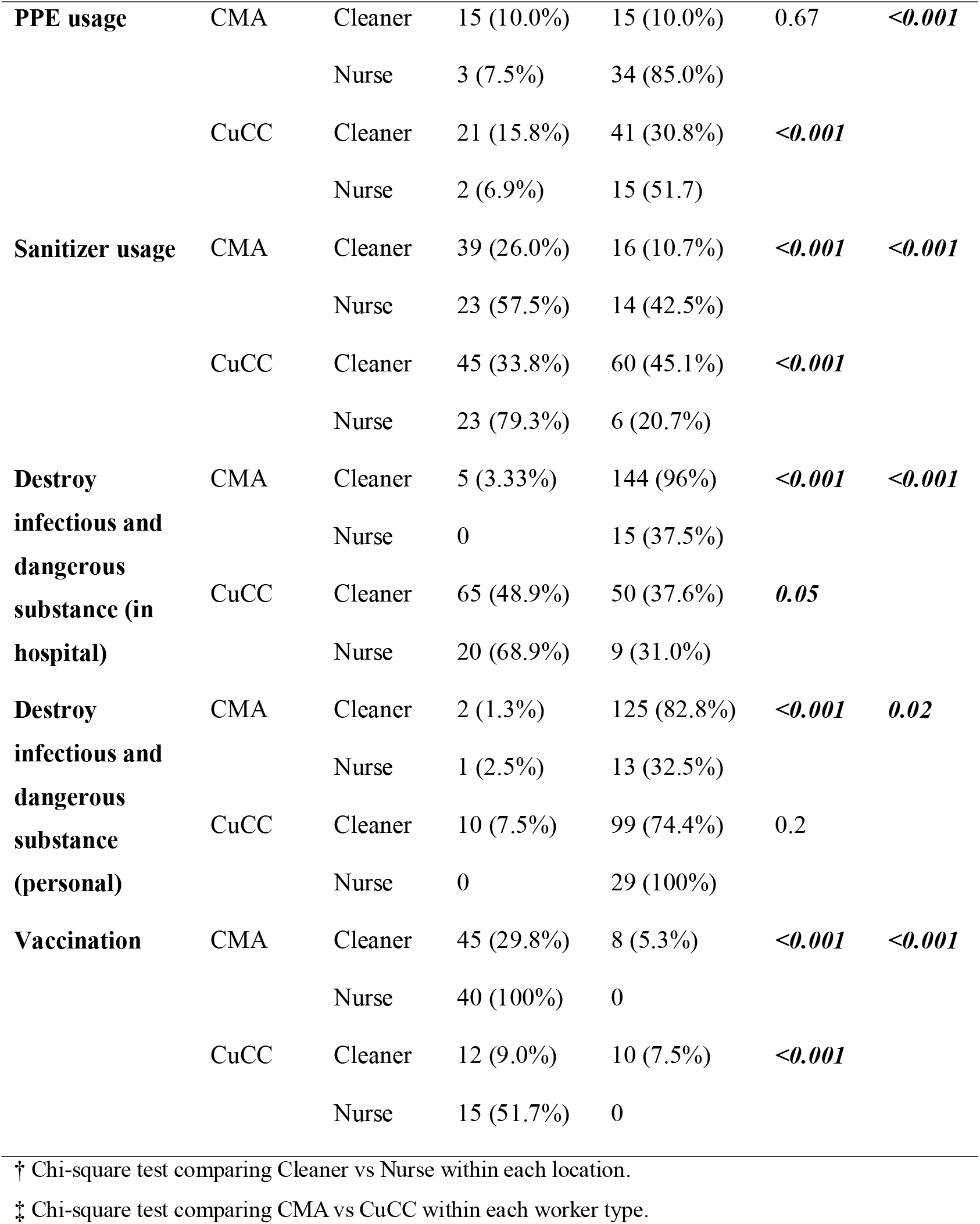
Personal hygiene practices regarding HCW.

**Figure 2.**
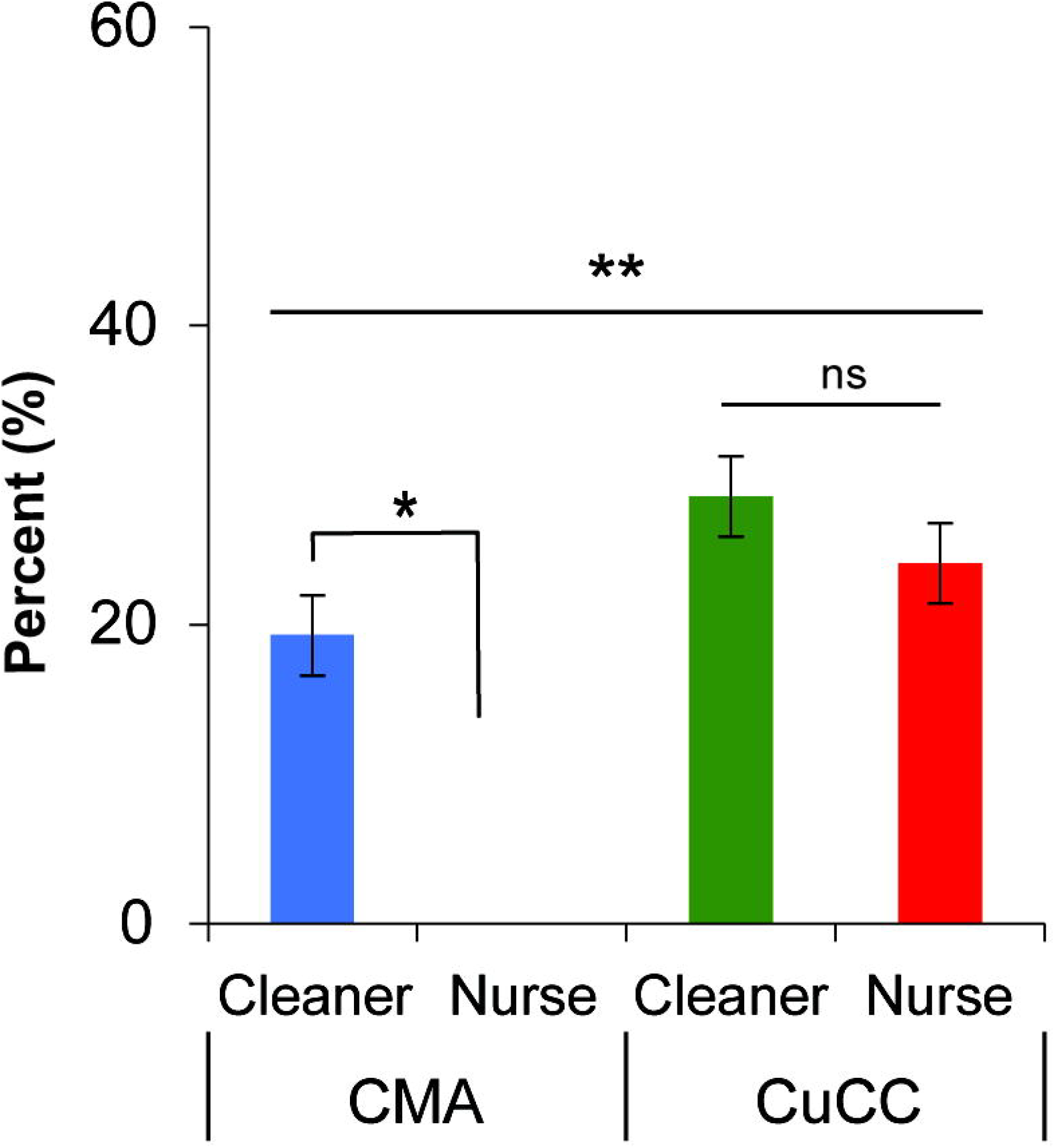
Practice of using separate color-coded bins for healthcare waste (HCW) management among cleaners and nurses at CMA and CuCC. Percentages indicate respondents reporting the use of separate color-coded bins for HCW segregation. Statistical significance was determined using the chi-square test: *P* < 0.05 for comparisons between cleaners and nurses within the same location (*), and between CMA and CuCC within the same worker category (**). “ns” indicates no statistically significant difference.

Protective practices varied considerably by role and location (Table 4). Nurses reported significantly higher rates of hand hygiene, PPE use, and vaccination than cleaners, particularly in CuCC (p < 0.001). Vaccination coverage was universal among nurses in CMA and substantially higher than among cleaners in both settings (p < 0.001). Hazardous waste destruction and monitoring practices were infrequently reported in CMA but were significantly more common in CuCC (p < 0.001).

### 3.5. Institutional support and monitoring

Access to institutional facilities and oversight differed markedly between professional groups (Table 5). Nurses were significantly more likely than cleaners to report access to changing rooms, health check-ups, and regular monitoring of HCW practices in both CMA and CuCC (p < 0.001). Monitoring systems were notably weaker in CMA, where fewer than 20% of participants reported routine oversight, compared with substantially higher reporting in CuCC (p < 0.01).

**Table 5.** Institutional support and monitoring practices regarding HCW management.

| Category | Location | Worker type | Yes, n (%) | *p*-value†<br>(Cleaner vs Nurse) | *p*-value‡<br>(Overall) |
| --- | --- | --- | --- | --- | --- |
| **Dedicated toilet facility for the HCW** | CMA | Cleaner | 56 (37.3%) | 0.62 | 0.79 |
|  |  | Nurse | 40 (100%) |  |  |
|  | CuCC | Cleaner | 44 (33.1%) | <0.001 |  |
|  |  | Nurse | 29 (100%) |  |  |
| **Dedicated changing room for the HCW** | CMA | Cleaner | 38 (25.3%) | <0.001 | 0.04 |
|  |  | Nurse | 38 (95.0%) |  |  |
|  | CuCC | Cleaner | 45 (33.8%) | <0.001 |  |
|  |  | Nurse | 22 (75.9%) |  |  |
| **Regular health check-up** | CMA | Cleaner | 13 (8.7%) | 0.44 | 0.007 |
|  |  | Nurse | 2 (5.0%) |  |  |
|  | CuCC | Cleaner | 11 (8.3%) | <0.001 |  |
|  |  | Nurse | 14 (48.8%) |  |  |
| **Regular monitoring** | CMA | Cleaner | 24 (16.0%) | 0.58 | 0.003 |
|  |  | Nurse | 5 (12.5%) |  |  |
|  | CuCC | Cleaner | 65 (48.9%) | 0.002 |  |
|  |  | Nurse | 23 (79.3%) |  |  |

† Chi-square test comparing Cleaner vs Nurse within each location.
‡ Chi-square or Fisher's exact test comparing CMA vs CuCC within each worker type.

## 4. Discussion

Effective healthcare waste (HCW) management is fundamental to infection prevention, occupational safety, and environmental protection, particularly in low- and middle-income countries (LMICs) where health systems face structural and resource constraints (Chartier, 2014). Using a knowledge–attitude–practice (KAP) framework, this study demonstrates a persistent disconnect between awareness and implementation of HCW management practices among nurses and cleaners in urban Bangladesh, consistent with reports from comparable settings (Chudasama et al, 2013; Islam et al., 2025).

### 4.1. Knowledge of healthcare workers on HCW management

Knowledge of HCW categories and color-coded segregation was uneven, with cleaners consistently demonstrating lower awareness than nurses. Similar professional disparities have been reported in hospitals across South Asia and Sub-Saharan Africa, reflecting differences in education, training access, and institutional prioritization of clinical staff (Motlatla et al., 2021; Leonard et al., 2022). However, higher knowledge among nurses particularly in CuCC did not translate into better compliance, reinforcing evidence that knowledge alone is insufficient to ensure safe HCW practices (Mathur et al., 2011).

Training coverage varied significantly by location and professional group, indicating the absence of standardized HCW training policies. Previous studies have shown that irregular, one-time, or theory-based training has limited long-term impact unless reinforced through supervision and monitoring (Felek et al., 2025; Chelly et al., 2024; Conti et al., 2024). The observed persistence of unsafe practices despite training supports this conclusion.

### 4.2. Attitudes and the knowledge–practice gap

Attitudes toward hygiene and environmental risk were overwhelmingly positive, particularly among nurses, mirroring findings from multiple LMIC hospital settings (Mathur et al., 2011; Hakim et al., 2020). High willingness to participate in further training suggests receptiveness to institutional interventions. Nevertheless, the pronounced gap between attitudes and actual practices highlights the critical role of enabling systems such as infrastructure availability, workload management, and accountability mechanisms in shaping behavior (Haylie et al., 2025; Affordofe et al., 2025; Tantum et al., 2025).

Lower environmental risk awareness among cleaners suggests that current training programs insufficiently address ecological and public health consequences of improper HCW disposal. Integrating environmental health perspectives into HCW training has been shown to improve risk perception and compliance, particularly among non-clinical staff (Savio et al., 2025; Thakan et al., 2025., Conti et al., 2024).

### 4.3. Practices and institutional determinants

Compliance with color-coded waste segregation—central to HCW management—was alarmingly low, particularly in CMA. Similar failures in segregation at source have been widely documented and are frequently attributed to inadequate bin availability, poor placement, lack of supervision, and weak enforcement rather than individual negligence (Chartier, 2014). The low compliance among nurses despite higher knowledge further underscores the dominance of structural barriers.

Occupational safety practices, including PPE use, hand hygiene, and vaccination, were markedly better among nurses than cleaners and were strongest in CuCC, where institutional support appeared more robust. Disparities in occupational protection for cleaners are well documented and raise serious ethical and public health concerns, given their direct exposure to hazardous waste (Ghebreyesus, 2022; Alam et al., 2022).

### 4.4. Institutional support and governance

Institutional support emerged as the most decisive determinant of HCW management performance. Facilities with better access to hygiene infrastructure, changing rooms, health monitoring, and routine supervision particularly in CuCC demonstrated relatively improved compliance. These findings align with evidence that governance, monitoring, and enforcement are stronger predictors of HCW management success than individual knowledge or attitudes (Chartier, 2014; Tilahun et al., 2023; Mutekanga et al., 2022).

## Conclusion

Consistent with global evidence, this study demonstrates that unsafe HCW management persists not due to lack of awareness, but because of systemic institutional failures. Addressing HCW risks in Bangladesh requires standardized training, equitable occupational protection for cleaners, adequate infrastructure, and enforceable monitoring systems, in line with WHO recommendations.

## Data Availability

The datasets generated and/or analyzed during the current study are available from the corresponding author upon reasonable request. Data are not publicly available due to institutional and participant confidentiality considerations.

## Author Contributions

**NNB, KNB, SA**: Investigation, Methodology, Data curation, Visualization, Validation, Writing – original draft; **RSRB, TMR, MAH**: Conceptualization, Project administration, Resources, Supervision, Writing – review & editing.

